# Predictive and analysis of COVID-19 cases cumulative total: ARIMA model based on machine learning

**DOI:** 10.1101/2022.01.24.22269791

**Authors:** Zehui Yan, Yanding Wang, Meitao Yang, Zhiqiang Li, Xinran Gong, Di Wu, Wenyi Zhang, Yong Wang

**Affiliations:** Department of Child and Adolescent Health, School of Public Health, China Medical University, Shenyang, 110122, China; Department of Epidemiology and Statistics, School of Public Health, China Medical University, Shenyang, 110122, China; Chinese PLA Center for Disease Control and Prevention, 100071, China

## Abstract

At present, COVID-19 poses a serious threat to global human health, and the cumulative confirmed cases in America, Brazil and India continue to grow rapidly. Therefore, the prediction models of cumulative confirmed cases in America, Brazil and India from August 1, 2021 to December 31, 2021 were established. In this study, the prevalence data of COVID-19 from 1 August 2021 to 31 December 2021 were collected from the World Health Organization website. Several ARIMA models were formulated with different ARIMA parameters. ARIMA (7,2,0), ARIMA (3,2,1), and ARIMA (10,2,4) models were selected as the best models for America, Brazil, and India, respectively. Initial combinations of model parameters were selected using the automated ARIMA model, and the optimized model parameters were then found based on Bayesian information criterion (BIC). The analytical tools autocorrelation function (ACF), and partial autocorrelation function (PACF) were used to evaluate the reliability of the model. The performance of different models in predicting confirmed cases from January 1, 2022 to January 5, 2022 was compared by using root mean square error (RMSE), mean absolute error (MAE), and mean absolute percentage error (MAPE). This study shows that ARIMA models are suitable for predicting the prevalence of COVID-19 in the future. The results of the analysis can shed light on understanding the trends of the outbreak and give an idea of the epidemiological stage of these regions. Besides, the prediction of COVID-19 prevalence trends of America, Brazil, and India can help take precautions and policy formulation for this epidemic in other countries.

## 1 Background

With the widespread of the new Coronavirus (2019-nCoV), it has become a global disease. This new virus was later named as COVID-19, which is a kind of respiratory infectious disease with lung inflammation. Its clinical symptoms mainly include respiratory symptoms such as dyspnea and respiratory distress syndrome, because of its similarity to the SARS virus so it is also called (SARS-CoV-2) [1].COVID-19 shows more special transmission characteristics than previous infectious diseases, which leads to its faster transmission speed, wider transmission range, higher transmission risk and rapid epidemic spread, posing a great threat to global public health security [2].

Since the discovery of COVID-19 cases in Wuhan, Hubei Province in December 2019, the epidemic has spread rapidly to the whole country. As of 26 December, over 278 million cases and just under 5.4 million deaths have been reported globally [3]. At present, the spread of pandemic in most countries is still growing and has not been effectively controlled. Since the novel coronavirus pneumonia in America, India, Brazil and other countries, the cumulative number of patients has been high. Therefore, the prevention and control of COVID-19 epidemic not only provides a more important basis for the construction of mathematical model of COVID-19 epidemic trend in most countries, but also provides decision-making basis for the prevention and control of global COVID-19 epidemic. It also provides a methodological basis for the establishment of prediction models for the prevention and control of other major infectious diseases.

In recent studies, different models have been used to predict COVID-19 prevalence, and mortality rate. For example, multiple linear regression [4], grey prediction model [5], simulation model [6], Holt model [7], LSTM model [8]. However, the spread of epidemic disease is random and will be affected by many factors. Therefore, the above statistical model can not accurately analyze the diffusion trend of epidemic and predict its future trend, and it is difficult to popularize the model.

The Automatic Regressive Integrated Moving Average (ARIMA) model has many advantages, such as simple structure and rapid applicability. The ARIMA model was successfully applied to the prediction and estimation of many prevalent diseases, such as typhoid fever [9], tuberculosis [10], influenza [11], brucellosis [12] and COVID-19 [13]. Since ARIMA methods do not contain much much mathematics or statistics, but also are capable of correlating regulation with short-term changing trends in the time series. So, the model is more suitable for the prediction analysis of epidemic diseases in the short term.

As of December 28, 2021, the cumulative prevalence of new coronal pneumonia was greatest in the United States, followed by India, Brazil. Due to the fastest spread of the new coronavirus, and the number of confirmed cases in the aforementioned countries is still continuously increasing. Therefore, it is of great significance to analyze the situation of new crown pneumonia at present and predict the epidemic trend.

The aim of this study is to estimate the cumulative cases of COVID-19 in America, India, and Brazil. The data analyzed in this study correspond to the period between 1 August 2021 and 31 December 2021. The data set was used to perform and analyze a case estimation model by applying different ARIMA models. Thus, in addition to enlightening the characteristics of the spread of the epidemic, it was aimed to provide authorities with realistic estimates for the peak time and intensity of the epidemic using models based on simple quantitative models. These models can help predict the health infrastructure and material needs that patients will need in these countries in the near future.

## 2. Methods

### 2.1 Data collection

This article is based on the official WHO website, and MS Excel was used to build a time-series database [14]. To create a stable and effective ARIMA model, at least 30 observations are required [15]. The new COVID-19 statistics released, selected daily confirmed cases of COVID-19 in three countries, America, India, and Brazil, as of August 1, 2021 - December 31, 2021, were used to construct a disease prediction model. and to forecast and evaluate the model prediction performance for confirmed case data for the next 5 day with 95% relative confidence intervals. (January 1, 2022 - January 5, 2022).

### 2.2 ARIMA model

ARIMA is a type of algorithm for the analysis and forecasting of time series data, namely the Box - Jenkin model, first proposed by Box and Jenkins in the 1970s [15].The ARIMA (p, d, q) model, known as the differential autoregressive moving average model.The modeling idea of the ARIMA model is to apply mathematical models to non-stationary time series After smoothing the data, it is used to estimate and extrapolate the state of something at some point in the future by analyzing the pattern of historical data and making future predictions based on that pattern and historical data from the past and the present. The cumulative number of confirmed cases of New Coronary Pneumonia is a random series, so the model can be considered as suitable for forecasting. Arima simulates and estimates the state of something at some point in the future. The ARIMA model includes the following steps. Step 1: Assessment of the model. Step 2: The model parameters were estimated.Step 3: Check the hypotheses of the model validation. Step 4: Modeling predictions. The structure of the ARIMA (p, d, q) model is Eq. (1).

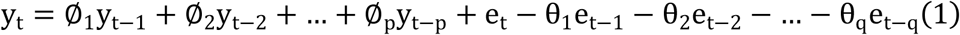

In Eq. (1),ϕ_*a*_(a=1,2,…,p) and ϕ_*b*_(b=0,1,2,…,q) are parameters of the model. y_*t*_ and ε_*t*_ represent the original value and arbitrary error at time step t.The arbitrary error represented by ε_*t*_ represents σ^2^ with zero mean and standard deviation. Taking the value q=0 in Eq. (1) works as AR model with order p and for p=0 it becomes the MA model with q order. So (p, q) are both important factors to determine ARIMA model.

### 2.3 Performance indices

In the study, we applied two performance evaluation indicators, namely RMSE, MAE and MAPE,were applied to test the predictive accuracy of the developed models.The formulations of these criteria are expressed Eq. (2) to Eq. (4), respectively [16].

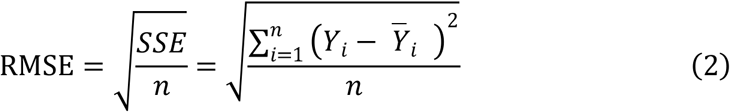

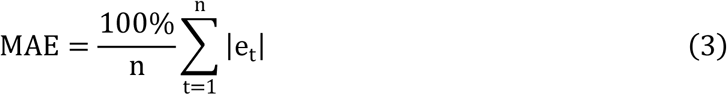

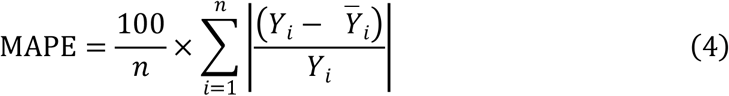

Where *y*_*t*_ is the observed value at time *t*, and *e*_*t*_ is also the difference between the observed and estimated value. In addition, *n* is the number of time points. Therefore, the RMSE and MAPE values can be used to indicate the fitting effect of the data. Lower RMSE, MAE, and MAPE values indicate a better fit of the data. All analyses were performed using SAS 9.4. software with a statistically significant level of *p* < 0.05.

## 3 Results and discussion

The aim of this paper is to perform a statistical, observational and predictive analysis of a cumulative confirmed case dataset from COVID-19 in three countries (America, Brazil, and India) by ARIMA model. The COVID-19 data set is divided into 153 days of training samples and the next 5-day of prediction samples. All statistical procedures were performed on the transformed COVID-19 data.

The first step of the ARIMA model is to control whether the time series is stationary and seasonal. A time series is considered as stationary if its statistical properties such as mean, variance, autocorrelation are constant over time, and it will make it easier to get accurate estimates [17]. However, because the prevalence of COVID-19 is unstable. Even after the first difference, it seems that the trends of all series not eliminated, so the second-order differences should be taken in the process of ARIMA test model, Shown in Table 2. The P values related to parameters are all less than 0.05, which proves to be white noise sequence. Autocorrelation Function (ACF), and Partial Autocorrelation Function (PACF) graphs were constructed to check the seasonality and stationarity. Estimated autocorrelations for the time series of America, Brazil, and India are shown in Fig. 1.

**Table 1.** Descriptive statistics on the prevalence of COVID-19 in America, India, and Brazil.

| Country | Mean | SE Mean | St. Dev | Minimum | Maximum | Skewness | Kurtosis |
| --- | --- | --- | --- | --- | --- | --- | --- |
| America | 43848993 | 391400 | 4841353 | 34798057 | 53506483 | -0.198 | -0.898 |
| India | 33746197 | 75158 | 929660 | 31655824 | 34838804 | -0.756 | -0.691 |
| Brazil | 21426569 | 55688 | 688818 | 19880273 | 22263834 | -0.649 | -0.771 |

**Table 2.** White noise autocorrelation check in America, Brazil and India.

| Country | Lag value | Chi square value | Variance | Pr> Chi square value* | Autocorrelation |  |  |  |  |  |
| --- | --- | --- | --- | --- | --- | --- | --- | --- | --- | --- |
| America | 6 | 21.49 | 6 | 0.0015 | -0.144 | -0.263 | 0.001 | 0.018 | -0.198 | -0.092 |
|  | 12 | 104.45 | 12 | <.0001 | 0.650 | -0.149 | -0.204 | 0.055 | -0.025 | -0.159 |
|  | 18 | 164.48 | 18 | <.0001 | -0.061 | 0.542 | -0.126 | -0.199 | 0.041 | -0.026 |
|  | 24 | 215.77 | 24 | <.0001 | -0.154 | -0.013 | 0.464 | -0.128 | -0.178 | 0.035 |
| Brazil | 6 | 49.69 | 6 | <.0001 | -0.487 | 0.106 | -0.220 | 0.120 | -0.093 | 0.034 |
|  | 12 | 54.22 | 12 | <.0001 | 0.109 | -0.052 | 0.062 | -0.091 | -0.036 | -0.001 |
|  | 18 | 55.83 | 18 | <.0001 | 0.013 | 0.085 | 0.004 | -0.046 | 0.002 | -0.002 |
|  | 24 | 58.81 | 24 | <.0001 | -0.065 | -0.001 | 0.099 | 0.010 | -0.005 | -0.051 |
| India | 6 | 22.85 | 6 | 0.0008 | -0.025 | -0.204 | -0.164 | -0.106 | -0.255 | 0.018 |
|  | 12 | 113.45 | 12 | <.0001 | 0.652 | 0.063 | -0.240 | -0.100 | -0.220 | -0.130 |
|  | 18 | 187.39 | 18 | <.0001 | -0.034 | 0.591 | 0.064 | -0.194 | -0.158 | -0.141 |
|  | 24 | 245.42 | 24 | <.0001 | -0.149 | 0.009 | 0.487 | 0.142 | -0.214 | -0.028 |

**Fig. 1.**
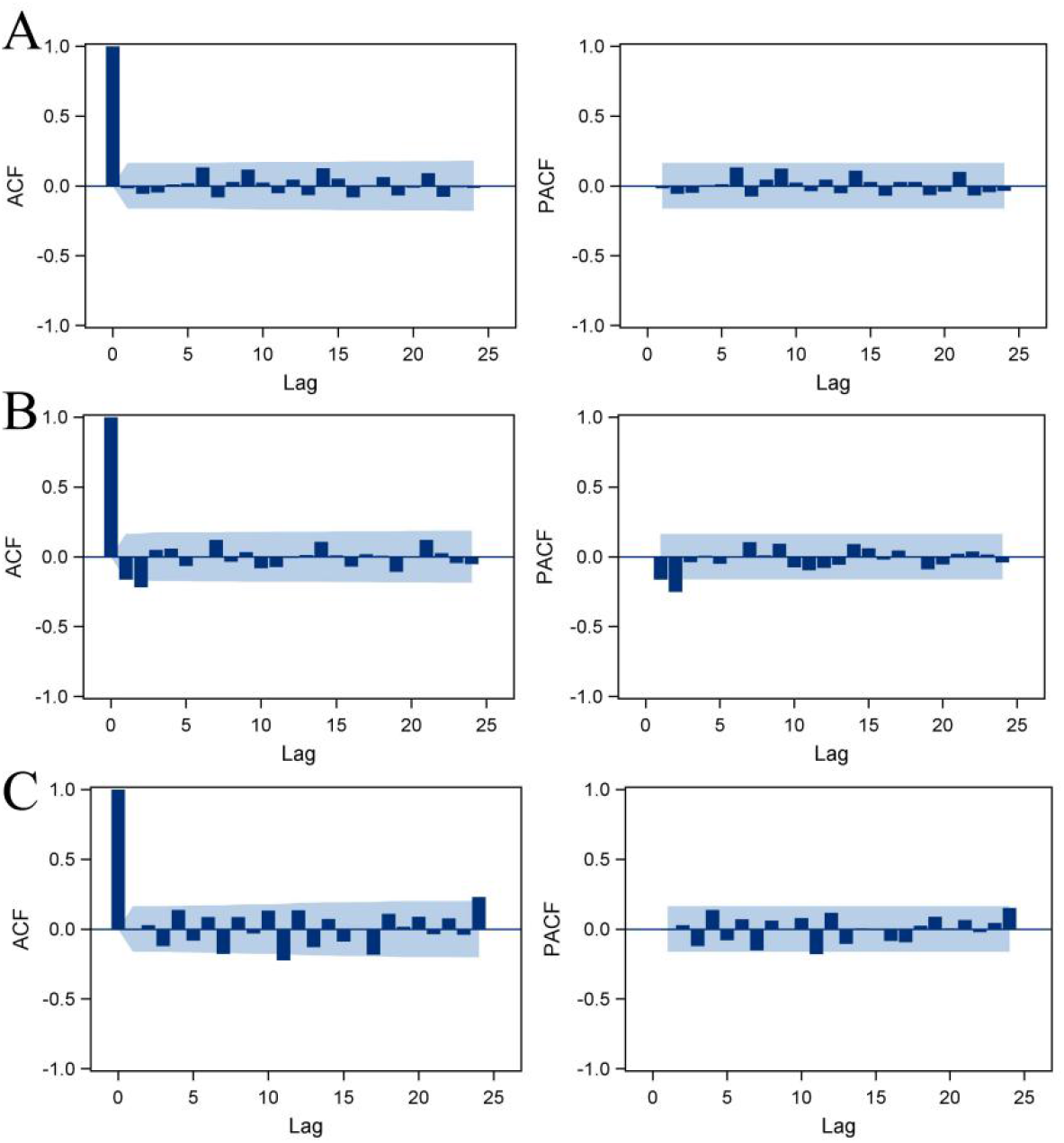
The estimated ACF and PACF graphs to predict the epidemiological trend of COVID-19 prevalence for (A) America, (B) Brazil, and (C) India.

Furthermore, according to the normality test, the model successfully achieves the independence of normal variables. It is further explained that the model is a normal conclusion and independent conclusion, as shown in Fig. 2. Secondly, adopting the minimum information criterion, the ARIMA (7,2,0), ARIMA (3,2,1), and ARIMA (10,2,4) models were chosen as the best models for America, Brazil, and India, respectively.

**Fig. 2.**
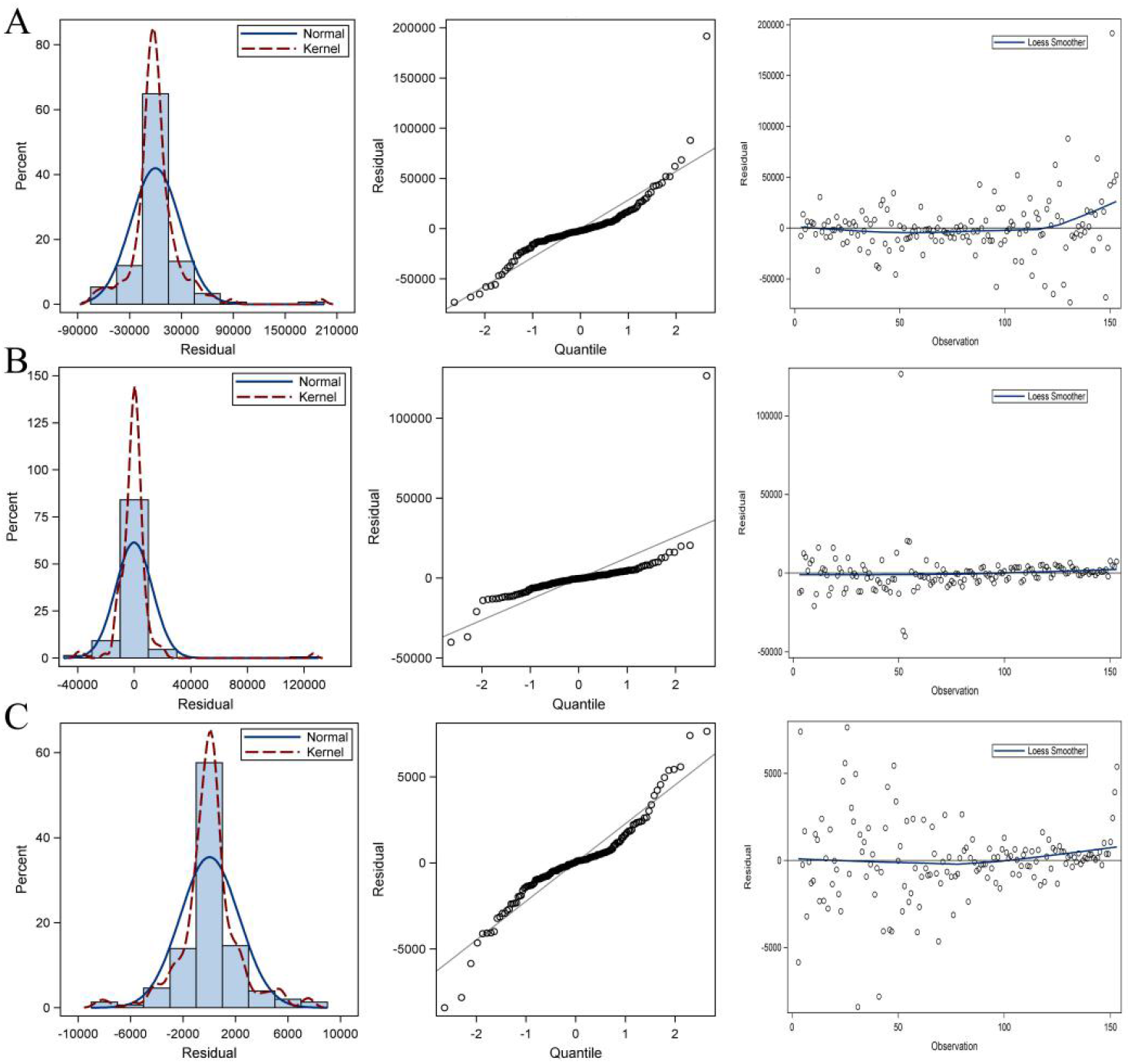
The normality test for (A) America, (B) Brazil, and (C) India.

The models fitted the COVID-19 data reasonably well (Table 3) with a minimum MAPE_*America*_ = 0.00132, MAPE_*Brazil*_ = 0.00048, and MAPE_*India*_ = 0.00021 values. Table 4 shows the parameter estimates for the best models. The p-values of the associated with the parameters are <0.05, so the terms are considerably different from zero at the 95.0% confidence level. Finally, the 5-day prediction results and trends of ARIMA model in this study are shown in Figure 3. And it can be seen that ARIMA has a good fitting degree at the early stage of prediction. And with the passage of prediction time, they all keep a stable effect.

**Table 3.** Comparison of tested ARIMA models.

| Country | RMSE | MAE | MAPE |
| --- | --- | --- | --- |
| America | 146766.8422 | 71442 | 0.00132 |
|  | 160155.0580 | 139506 | 0.00256 |
|  | 166469.8635 | 107320 | 0.00196 |
|  | 189234.0667 | 108349 | 0.00196 |
|  | 244703.0000 | 244703 | 0.00437 |
| Brazil | 27042.4575 | 10753 | 0.00048 |
|  | 29752.5022 | 20353 | 0.00091 |
|  | 32283.1273 | 25822 | 0.00116 |
|  | 35070.1101 | 28277 | 0.00127 |
|  | 40746.0000 | 40746 | 0.00183 |
| India | 62211.6553 | 7494 | 0.00021 |
|  | 69453.7441 | 20466 | 0.00059 |
|  | 79323.0381 | 40892 | 0.00117 |
|  | 92747.7155 | 66586 | 0.00190 |
|  | 113007.0000 | 113007 | 0.00323 |

**Table 4.** Parameters of ARIMA models.

| Country | Best model | Coefficient | Standart error | t-Statistic | p-Value |
| --- | --- | --- | --- | --- | --- |
| America | ARIMA (7,2,0) | 0.82314 | 0.08391 | 9.81 | <.001 |
| Brazil | ARIMA (3,2,1) | -0.28163 | 0.08313 | -3.39 | <.001 |
| India | ARIMA (10,2,4) | -1.20779 | 0.29914 | -4.04 | <.001 |

**Fig. 3.**
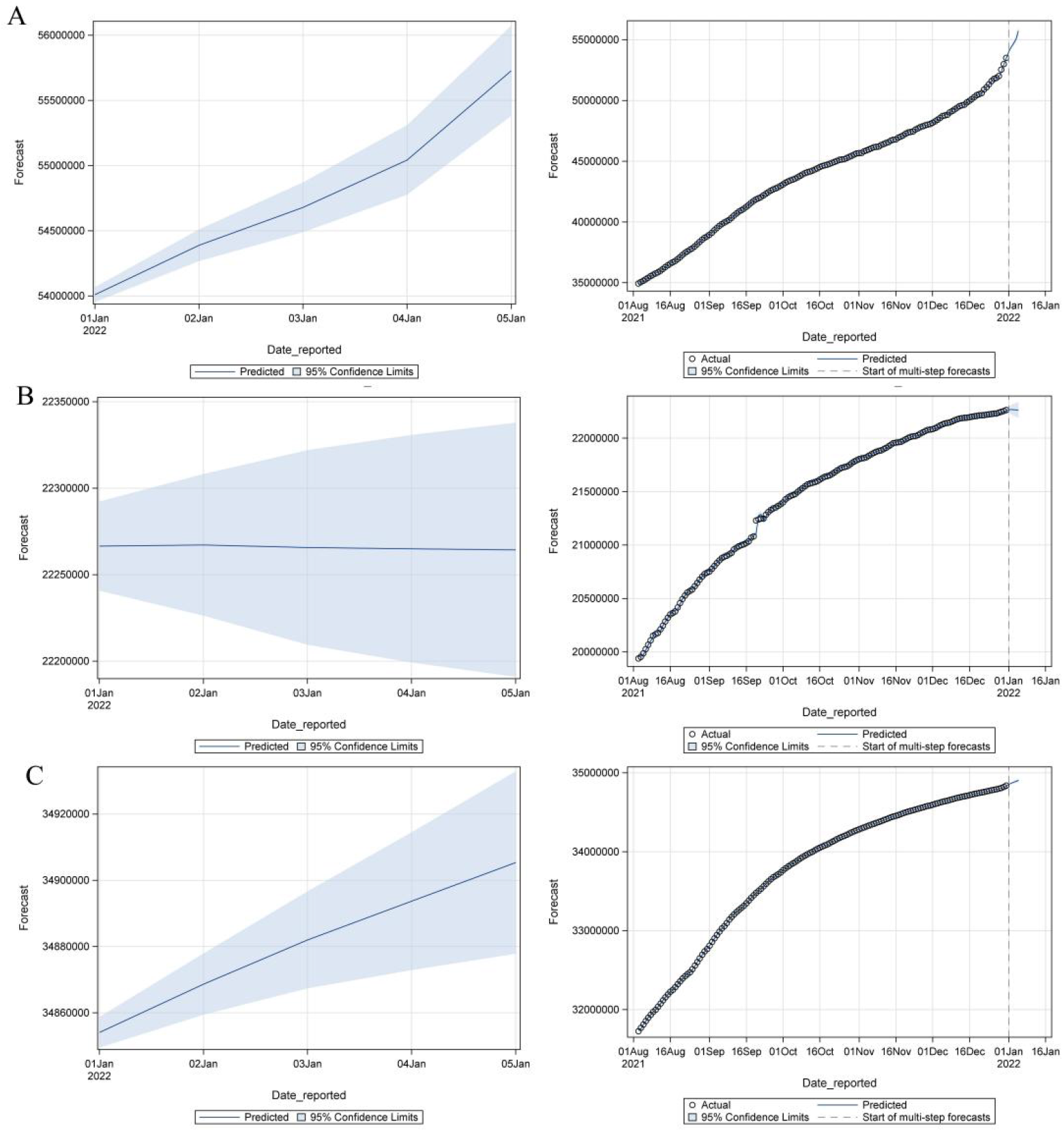
Time-series plots for (A) America, (B) Brazil, and (C) India.

As seen in Table 5, the next 5-day estimate of confirmed cases may be between 54009063–55727411 in America, 22266486–22264332 in Brazil, and 34854085–34905351 in India.

**Table 5.**
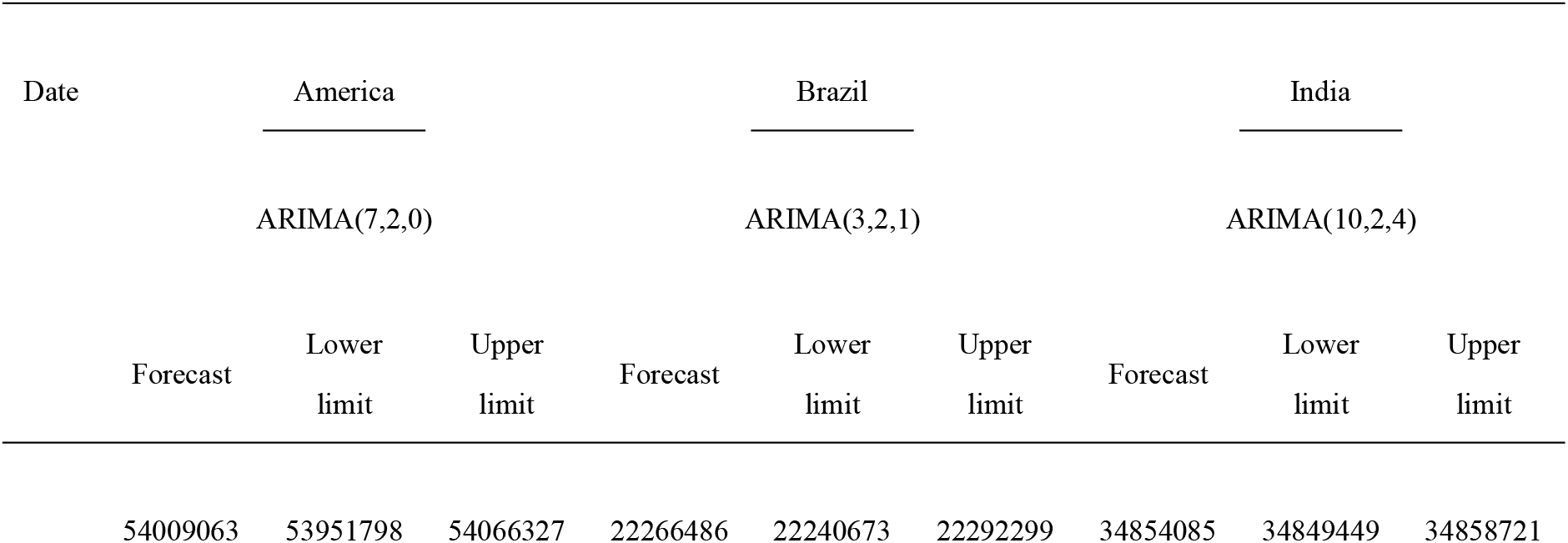
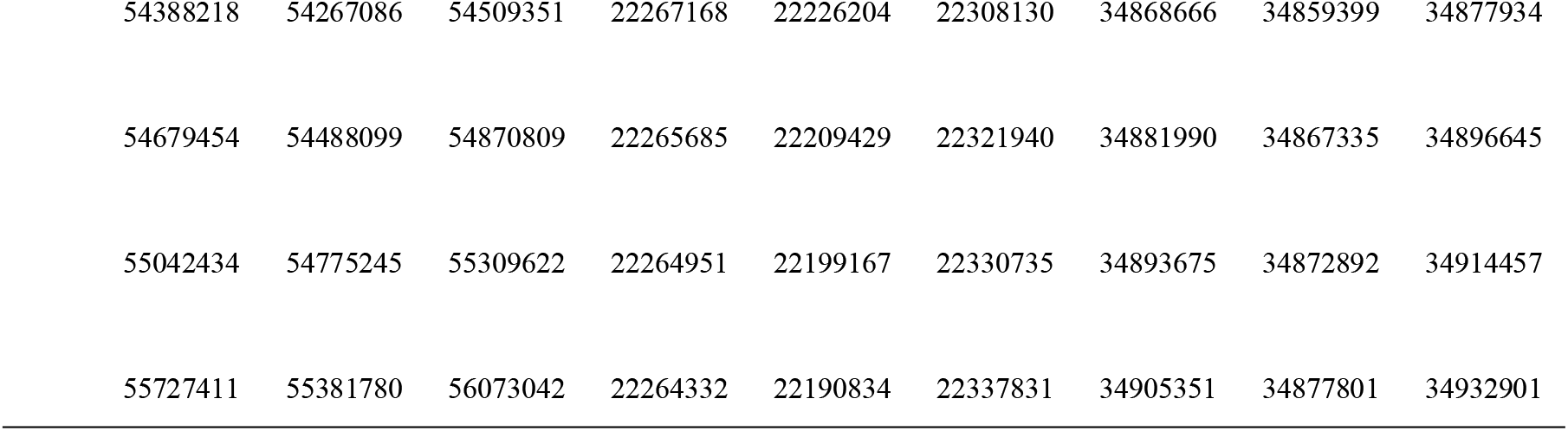
Prediction of total confirmed cases of COVID-19 for the next 5-day according to ARIMA models with 95% confidence interval.

In our research, according to the prediction results of the cumulative confirmed cases in COVID-19 of three countries, the prediction results for the next 5-day all showed better trends. Therefore,this study further proves that ARIMA models can be used for pandemic prediction, which is conducive to better planning and management of infectious diseases.

## 4 conclusion

COVID-19 is currently spreading worldwide, and the significance of the prediction and analysis of the outbreak is very important. Time series analysis is instrumental in developing hypotheses to understand the prevalence trend of various diseases and forecast the dynamics of observed phenomena, and then in the construction of a quality control system. ARIMA model is one of the most commonly used time series forecasting methods because of its simplicity and systematic structure and acceptable forecasting performance [18].

In this study, the current situation of the COVID-19 pandemic in America, Brazil, and India was presented, and the sustained trend and extent of the outbreak were estimated by the ARIMA model. Therefore, we further demonstrated that the ARIMA model can reliably and precise predictions of the onset of new crown outbreaks. That can help governments as a reference to decide on emergency macroeconomic strategies and medical resource allocation, regulation of production activities, and even for the national economic development of countries, and its stable prediction of COVID-19 provides more new ideas and reference value.

## Data Availability

According to the reasonable requirements of the author, all the data generated in this study can be obtained. All the data generated in the current work are included in the manuscript, and all the data generated can be consulted online.

https://www.who.int/emergencies/disease-outbreak-news

